# Long Noncoding RNA Associations Define an Interferon–Myeloid Immune Axis in Kawasaki Disease

**DOI:** 10.64898/2026.05.21.26353728

**Authors:** Fang Liu, Xing Xue, Zhi Han, Bo Jin, Weiwei Li, Naoto Ozawa, Takumi Ichikawa, Ellen Ling, Xinyang Zhao, Henry Chubb, Scott R. Ceresnak, Gary L. Darmstadt, Doff B. McElhinney, Harvey J. Cohen, Seda Tierney, Xuefeng B. Ling

**Affiliations:** Heart Center, Children’s Hospital of Fudan University, Shanghai, China; School of Medicine, Stanford University, Stanford, CA, USA; OncoOmicsDx Clinical Laboratory, Rockville, MD, USA; Nippon Life Insurance Company, Osaka, Japan; Florida State University, Tallahassee, FL 32306, USA; University of Kansas Medical Center, Kansas City, KS 66160, USA

## Abstract

Kawasaki disease (KD) is an acute pediatric vasculitis characterized by dysregulated host immune responses and risk of coronary artery injury. Although a two-transcript IFI27–MCEMP1 axis has been clinically validated to distinguish KD from other febrile illnesses, the long noncoding RNA (lncRNA) context of this interferon–myeloid imbalance remains incompletely understood. We evaluated whether peripheral blood mononuclear cell (PBMC)-derived lncRNAs are altered in KD and associated with the interferon and myeloid components of the IFI27–MCEMP1 transcriptomic axis. Children younger than 8 years with suspected KD were prospectively enrolled at the Children’s Hospital of Fudan University from 2024 to 2025. The newly enrolled cohort included 55 children with KD and 48 febrile controls. For integrated immune-transcript association analyses, these data were combined with two previously characterized same-site cohorts, yielding 188 children with KD and 175 febrile controls. Expression of IFI27, MCEMP1, CHROMR, MALAT1, and NEAT1 was measured by reverse transcription quantitative PCR and normalized to GAPDH using ΔCt values. In the newly enrolled cohort, the IFI27–MCEMP1 axis reproduced discrimination between KD and febrile controls, with an area under the receiver operating characteristic curve of 0.88; performance was similar in the integrated cohort, with an area under the curve of 0.89. In PBMC lncRNA analyses, CHROMR and MALAT1 ΔCt values were significantly higher in KD than in febrile controls, indicating lower relative expression, whereas NEAT1 did not show a significant KD-specific differential-expression signal. CHROMR showed the strongest association with the IFI27 interferon-associated component, while MALAT1 showed weaker but directionally informative associations with both IFI27 and MCEMP1, including an inverse association with MCEMP1. These findings support an lncRNA-associated interferon–myeloid immune architecture in KD, marked by coordinated attenuation of IFI27, CHROMR, and MALAT1 together with increased MCEMP1. This PBMC RNA pattern provides a biologically interpretable framework for KD immune dysregulation and generates testable hypotheses regarding RNA-regulatory programs in KD vasculitis.

## INTRODUCTION

Kawasaki disease (KD) is an acute systemic vasculitis of childhood and remains the leading cause of acquired heart disease in children in many developed countries [1; 2] Although timely treatment with intravenous immunoglobulin substantially reduces the risk of coronary artery aneurysms[3], the immunopathogenesis of KD remains incompletely understood. The disease is characterized by fever, mucocutaneous inflammation, systemic immune activation, and a predilection for medium-sized arteries, particularly the coronary arteries[4]. The acute phase of KD involves coordinated perturbation of innate and adaptive immune pathways, including myeloid-cell activation, cytokine signaling, endothelial inflammation, and vascular remodeling. However, the circulating immune-transcriptional programs that distinguish KD from other pediatric febrile illnesses remain only partially defined.

A major challenge in understanding KD biology is that affected children often present during an acute febrile inflammatory state that overlaps clinically and immunologically with viral or bacterial infections. Children with incomplete or evolving KD may lack sufficient clinical criteria at presentation, while febrile controls may show overlapping inflammatory features such as rash, conjunctival injection, mucosal changes, lymphadenopathy, elevated C-reactive protein, leukocytosis, or neutrophilia[5]. This overlap complicates clinical diagnosis, but it also creates an opportunity to interrogate disease-specific host-response patterns. Blood transcriptomic profiling can capture immune-state information that is not apparent from clinical signs or conventional inflammatory markers alone. In this context, transcriptomic signatures may serve not only as diagnostic adjuncts but also as windows into the immune architecture of KD.

We recently validated a parsimonious two-transcript IFI27–MCEMP1 reverse transcription quantitative polymerase chain reaction (RT-qPCR) axis that distinguishes KD from other pediatric febrile illnesses [6]. IFI27 encodes an interferon-stimulated gene associated with antiviral host-response programs [7] whereas MCEMP1 is linked to myeloid and inflammatory activation [8; 9]. The IFI27–MCEMP1 axis therefore captures a biologically interpretable contrast between interferon-associated and myeloid-associated transcriptional states. In KD, this pattern is characterized by relative attenuation of IFI27 and elevation of MCEMP1 compared with febrile controls, suggesting that KD is not simply a state of nonspecific inflammation but instead reflects a distinct immune-transcriptional configuration. The biological context of this interferon– myeloid imbalance, however, remains unresolved. In particular, it is unclear whether the IFI27–MCEMP1 axis represents an isolated downstream biomarker pattern or is embedded within broader RNA-regulatory programs relevant to KD vasculitis.

Long noncoding RNAs (lncRNAs) are increasingly recognized as regulators of immune-cell identity, inflammatory signaling, and cytokine-responsive transcription. Through chromatin remodeling, transcription-factor scaffolding, enhancer regulation, nuclear organization, RNA stability, and post-transcriptional control, lncRNAs can modulate the amplitude, timing, and specificity of immune responses. In inflammatory and antiviral contexts, lncRNAs may amplify or restrain interferon signaling, regulate myeloid-cell activation, and coordinate nuclear transcriptional responses to immune stress. These properties make lncRNAs plausible contributors to the RNA-regulatory landscape underlying KD immune dysregulation.

Several candidate lncRNAs are particularly relevant to interferon and myeloid biology. CHROMR, also known as cholesterol homeostasis regulator of macrophage response, has been implicated in macrophage immune responses and reported to interact with STAT1- and IRF1-associated transcriptional complexes, supporting interferon-stimulated gene expression [10]. NEAT1 regulates paraspeckle formation and has been linked to interferon-responsive transcriptional organization during inflammatory stress [11]. MALAT1 has been associated with regulation of inflammatory signaling, including NF-κB–related pathways in immune and vascular contexts [12]. These lncRNAs therefore provide biologically plausible candidates for evaluating whether the IFI27– MCEMP1 immune axis is accompanied by broader lncRNA perturbation in KD.

We hypothesized that selected PBMC-derived lncRNAs would be altered in children with KD and associated with the interferon and myeloid components of the IFI27– MCEMP1 transcriptomic axis. To test this hypothesis, we profiled CHROMR, MALAT1, and NEAT1 together with IFI27 and MCEMP1 in a newly enrolled cohort of children with KD and febrile controls, and integrated these data with two previously characterized same-site cohorts. We evaluated whether candidate lncRNAs differed between KD and febrile controls, whether their expression patterns were associated with IFI27 and MCEMP1, and whether these relationships supported an interpretable model of PBMC immune dysregulation in KD. By positioning the IFI27–MCEMP1 pair as an immune-axis anchor rather than solely as a diagnostic classifier, this study aimed to define an lncRNA-associated interferon–myeloid RNA architecture in KD and to generate mechanistic hypotheses for future studies of immune regulation, vascular inflammation, and disease endotyping in KD vasculitis.

## METHODS

### Study design and PBMC transcriptomic analysis cohorts

We conducted a diagnostic validation and biomarker association study in children evaluated for suspected Kawasaki disease (KD). The study was designed to address two related objectives. First, we assessed the reproducibility of a previously validated IFI27–MCEMP1 reverse transcription quantitative polymerase chain reaction (RT-qPCR) diagnostic score in a newly enrolled cohort of children with KD and febrile controls. Second, we evaluated whether selected peripheral blood mononuclear cell (PBMC)-derived long noncoding RNAs (lncRNAs) were differentially expressed in KD and associated with the interferon and myeloid components of the IFI27–MCEMP1 diagnostic axis.

Children younger than 8 years were prospectively enrolled at the Children’s Hospital of Fudan University, Shanghai, China, between 2024 and 2025. The newly enrolled validation cohort, referred to as batch 3, included 55 children with KD and 48 febrile controls. For integrated lncRNA and diagnostic-score analyses, batch 3 was combined with two previously characterized same-site cohorts generated using comparable enrollment criteria, sample-processing procedures, and RT-qPCR protocols. Batch 1 included 53 children with KD and 81 febrile controls, and batch 2 included 80 children with KD and 46 febrile controls. The integrated same-site cohort therefore included 188 children with KD and 175 febrile controls. Cohort assembly, sample numbers, and analytic use of each cohort are summarized in Table 1 and Figure 1.

**Table 1.** Cohort assembly and analytic samples.

| Cohort component | Source/role in present study | KD, No. | Febrile controls, No. | Total, No. | Primary use |
| --- | --- | --- | --- | --- | --- |
| Batch 1 | Previously characterized same-site cohort | 53 | 81 | 134 | Integrated lncRNA and diagnostic-score analyses |
| Batch 2 | Previously characterized same-site cohort | 80 | 46 | 126 | Integrated lncRNA and diagnostic-score analyses |
| Batch 3 | Newly enrolled validation cohort | 55 | 48 | 103 | Independent diagnostic validation and lncRNA profiling |
| Integrated same-site cohort | Batches 1–3 combined | 188 | 175 | 363 | Primary integrated lncRNA, correlation, and diagnostic-score analyses |

**Abbreviations:** FC, febrile control; KD, Kawasaki disease; lncRNA, long noncoding RNA; RT-qPCR, reverse transcription quantitative polymerase chain reaction. **Note:** Batch 3 represents the newly enrolled validation cohort. The integrated same-site cohort combines all three batches for lncRNA association analyses and diagnostic-score evaluation. PBMC RNA was profiled by RT-qPCR for IFI27, MCEMP1, CHROMR, MALAT1, NEAT1, and GAPDH.

**Figure 1.**
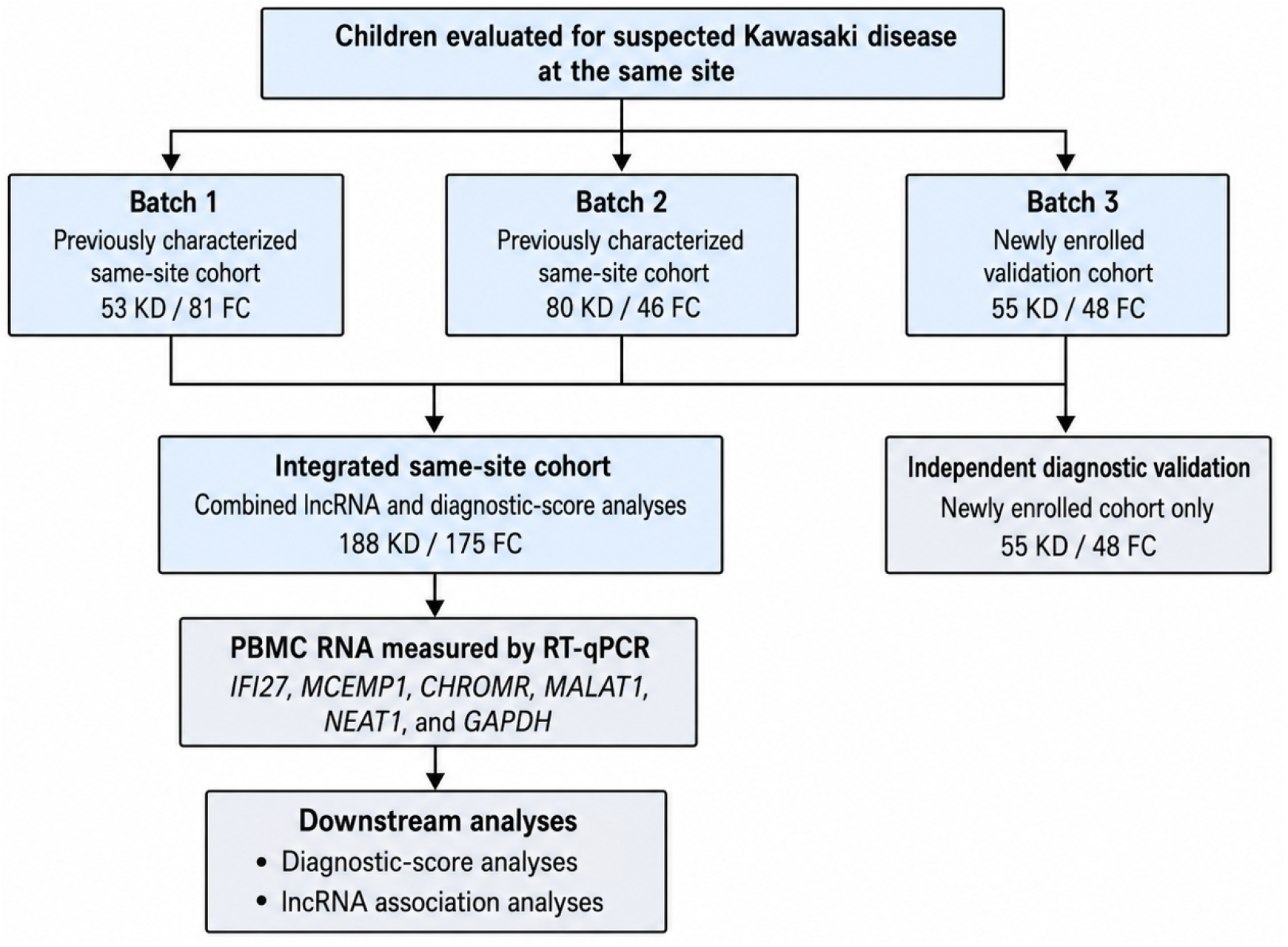
Study design and PBMC RNA profiling cohorts. Flow diagram showing assembly of the newly enrolled validation cohort and integrated same-site cohort used for diagnostic-score and long noncoding RNA analyses. Batch 3 comprised the newly enrolled validation cohort of 55 children with Kawasaki disease (KD) and 48 febrile controls (FC). Batches 1 and 2 were previously characterized same-site cohorts comprising 53 KD/81 FC and 80 KD/46 FC participants, respectively. Integration of all three batches yielded 188 KD and 175 FC participants for combined diagnostic-score and lncRNA association analyses. Peripheral blood mononuclear cell RNA was analyzed by RT-qPCR for IFI27, MCEMP1, CHROMR, MALAT1, NEAT1, and GAPDH. **Abbreviations:** FC, febrile control; IFI27, interferon alpha-inducible protein 27; KD, Kawasaki disease; lncRNA, long noncoding RNA; MCEMP1, mast cell-expressed membrane protein 1; PBMC, peripheral blood mononuclear cell; RT-qPCR, reverse transcription quantitative polymerase chain reaction.

### Participants and clinical classification

Eligible participants were children presenting with fever and clinical concern for KD or children evaluated for alternative febrile illnesses. KD was diagnosed according to the 2017 American Heart Association criteria for complete or incomplete KD. Children were classified as having KD on the basis of clinical features, laboratory findings, and echocardiographic evaluation. Complete KD was defined by fever with sufficient principal clinical criteria, whereas incomplete KD was diagnosed when children did not fulfill complete clinical criteria but met guideline-supported clinical, laboratory, or echocardiographic criteria for KD.

Febrile controls were children evaluated for alternative febrile illnesses who did not meet diagnostic criteria for KD. Febrile-control diagnoses were grouped as viral febrile illness, bacterial febrile illness, or other inflammatory febrile illness. Diagnostic classification of febrile controls was based on clinical assessment, laboratory testing, microbiologic studies when available, and treating-physician adjudication. Children were excluded from the analytic cohort if clinical data were insufficient for diagnostic classification or if RNA quantity or quality was inadequate for RT-qPCR analysis.

Demographic, clinical, and laboratory variables were abstracted from the clinical record. These included age, sex, fever duration at sample collection, hemoglobin concentration, C-reactive protein concentration, platelet count, white blood cell count, incomplete KD status, coronary artery status, and febrile-control diagnostic subgroup. Clinical and laboratory characteristics of the newly enrolled validation cohort are reported in Table 2.

**Table 2.**
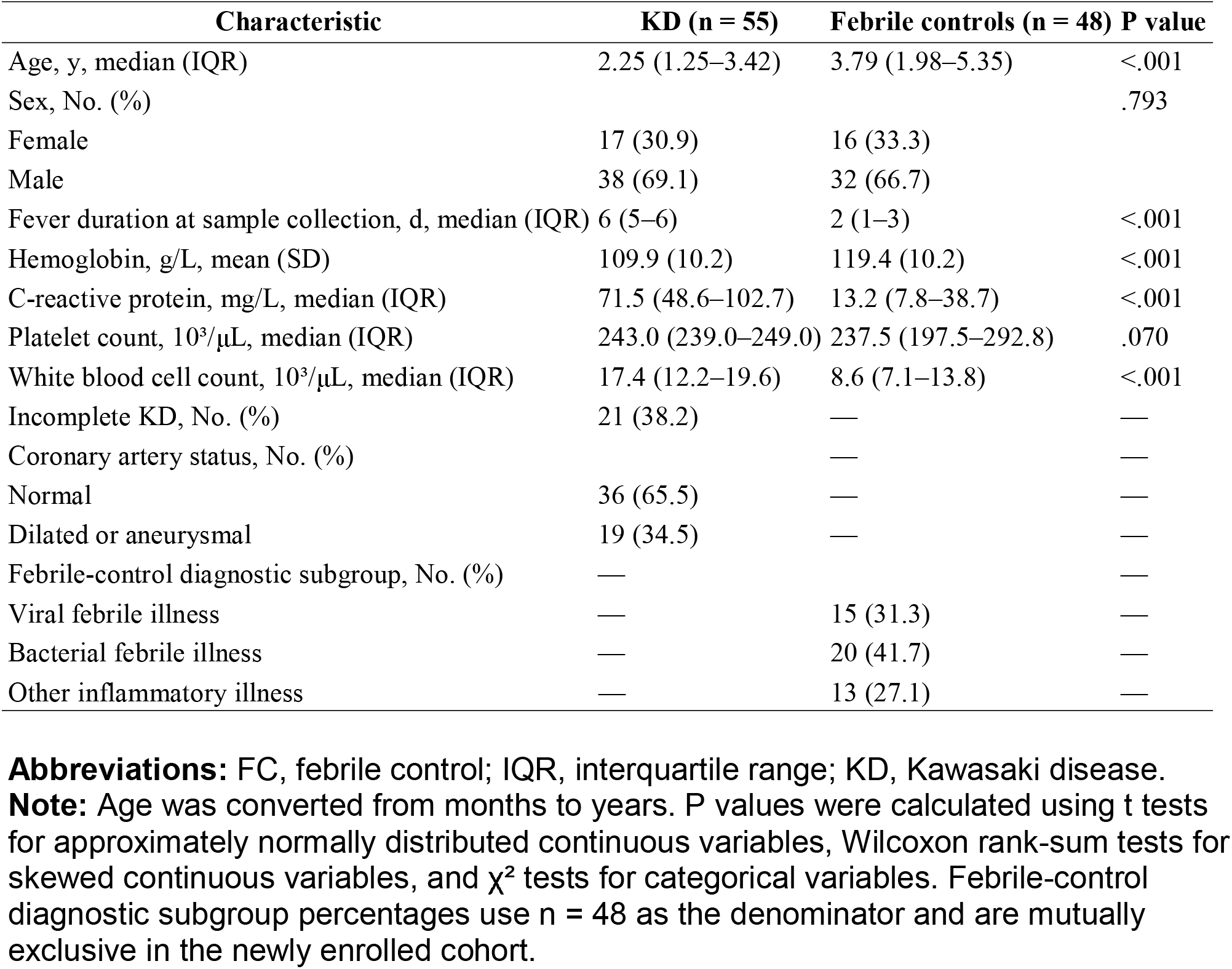
Demographic and clinical features of the newly enrolled cohort.

### Ethics approval and consent

The study was approved by the institutional review board of the Children’s Hospital of Fudan University. Written informed consent was obtained from the parents or legal guardians of all participants before study procedures. The study was conducted in accordance with applicable ethical standards for human participant research.

### Blood processing, RNA extraction, and RT-qPCR

Peripheral blood samples were obtained during the acute febrile illness and before intravenous immunoglobulin treatment in children diagnosed with KD. PBMCs were isolated from peripheral blood using standardized laboratory procedures and processed for total RNA extraction. RNA quantity and purity were assessed spectrophotometrically, and RNA quality was verified before reverse transcription. Complementary DNA was synthesized from total RNA using random hexamer priming.

Expression of IFI27, MCEMP1, CHROMR, MALAT1, NEAT1, and GAPDH was measured by RT-qPCR. qPCR reactions were performed in duplicate using SYBR Green chemistry on a real-time PCR platform. GAPDH was used as the endogenous reference gene for normalization. No-template controls were included to monitor contamination, and melt-curve analysis was used to confirm amplification specificity. Primer sequences, amplicon sizes, amplification efficiencies, and linearity metrics for GAPDH, CHROMR, NEAT1, and MALAT1 are provided in Table 3. IFI27 and MCEMP1 assay details were based on the companion clinically validated diagnostic assay report.

**Table 3.**
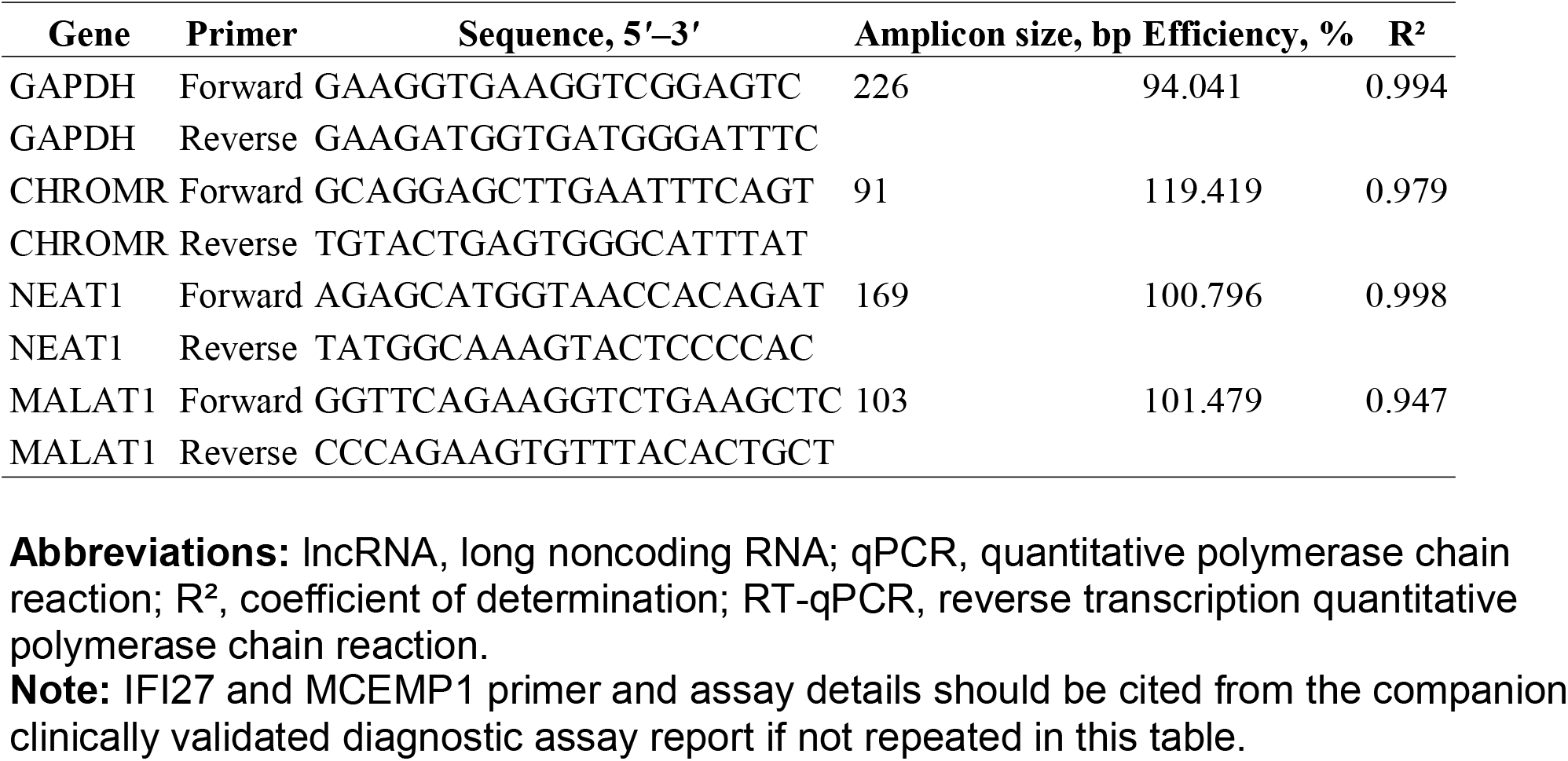
RT-qPCR primer sequences and assay parameters for lncRNA profiling.

### Transcript normalization and interferon–myeloid score definition

Gene expression was quantified using cycle threshold values normalized to GAPDH. For each target transcript, ΔCt was calculated as:

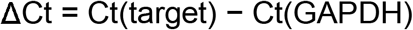

Higher ΔCt values indicate lower relative expression. The prespecified IFI27–MCEMP1 KD diagnostic score was calculated as:

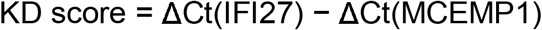

This score reflects the reciprocal expression pattern observed in KD, in which IFI27 expression is relatively lower and MCEMP1 expression is relatively higher compared with many other pediatric febrile illnesses. The diagnostic-positive operating point was KD score ≥2.5. This threshold was applied without recalibration in the newly enrolled validation cohort and in the integrated same-site cohort. No model refitting, coefficient adjustment, or post hoc threshold optimization was performed.

Diagnostic performance of the IFI27–MCEMP1 score was evaluated in the newly enrolled cohort and in the integrated same-site cohort. The corresponding diagnostic-performance estimates are reported in Table 4, and score distributions with receiver operating characteristic analyses are shown in Figure 2.

**Table 4.** Reproducibility of the IFI27–MCEMP1 interferon–myeloid score across febrile-control comparators.

| Cohort | Comparator | n | AUC (95% CI) | Sensitivity, % (95% CI) | Specificity, % (95% CI) | PPV, % (95% CI) | NPV, % (95% CI) |
| --- | --- | --- | --- | --- | --- | --- | --- |
| Newly enrolled cohort | All FC | 55 KD / 48 FC | 0.88 (0.80–0.95) | 81.8 (69.1–90.9) | 87.5 (74.8–95.3) | 88.2 (76.1–95.6) | 80.8 (67.5–90.4) |
| Integrated same-site cohort | All FC | 188 KD / 175 FC | 0.89 (0.86–0.93) | 85.6 (79.8–90.3) | 84.6 (78.4–89.6) | 85.6 (79.8–90.3) | 84.6 (78.4–89.6) |
| Newly enrolled cohort | Viral FC | 55 KD / 15 FC | 0.97 (0.93–1.00) | 81.8 (69.1–90.9) | 100.0 (78.2–100.0) | — | — |
| Newly enrolled cohort | Bacterial FC | 55 KD / 20 FC | 0.79 (0.65–0.93) | 81.8 (69.1–90.9) | 75.0 (50.9–91.3) | — | — |
| Newly enrolled cohort | Other inflammatory FC | 55 KD / 13 FC | 0.90 (0.79–1.00) | 81.8 (69.1–90.9) | 92.3 (64.0–99.8) | — | — |
| Integrated same-site cohort | Viral FC | 188 KD / 62 FC | 0.96 (0.93–0.98) | 85.6 (79.8–90.3) | 95.2 (86.5–99.0) | — | — |
| Integrated same-site cohort | Bacterial FC | 188 KD / 60 FC | 0.81 (0.73–0.88) | 85.6 (79.8–90.3) | 75.0 (62.1–85.3) | — | — |
| Integrated same-site cohort | Other inflammatory FC | 188 KD / 55 FC | 0.92 (0.87–0.96) | 85.6 (79.8–90.3) | 83.6 (71.2–92.2) | — | — |

**Abbreviations:** AUC, area under the receiver operating characteristic curve; FC, febrile control; KD, Kawasaki disease; NPV, negative predictive value; PPV, positive predictive value.
**Note:** KD score = $\Delta\text{Ct}(\text{IFI27}) - \Delta\text{Ct}(\text{MCEMP1})$ . The diagnostic-positive operating point was KD score $\geq 2.5$ . Sensitivity is repeated across comparator-specific rows because the same KD cases were compared with each FC subgroup within each cohort. PPV and NPV are reported only for the full FC comparison because predictive values depend on the case-control composition and may not generalize to clinical prevalence. AUC 95% CIs were estimated by the DeLong method; CIs for sensitivity, specificity, PPV, and NPV used exact binomial methods. Historical integrated FC subgroup annotations include two non-mutually exclusive febrile-control labels; therefore comparator-specific analyses should be interpreted as subgroup contrasts rather than mutually exclusive prevalence categories.

**Figure 2.**
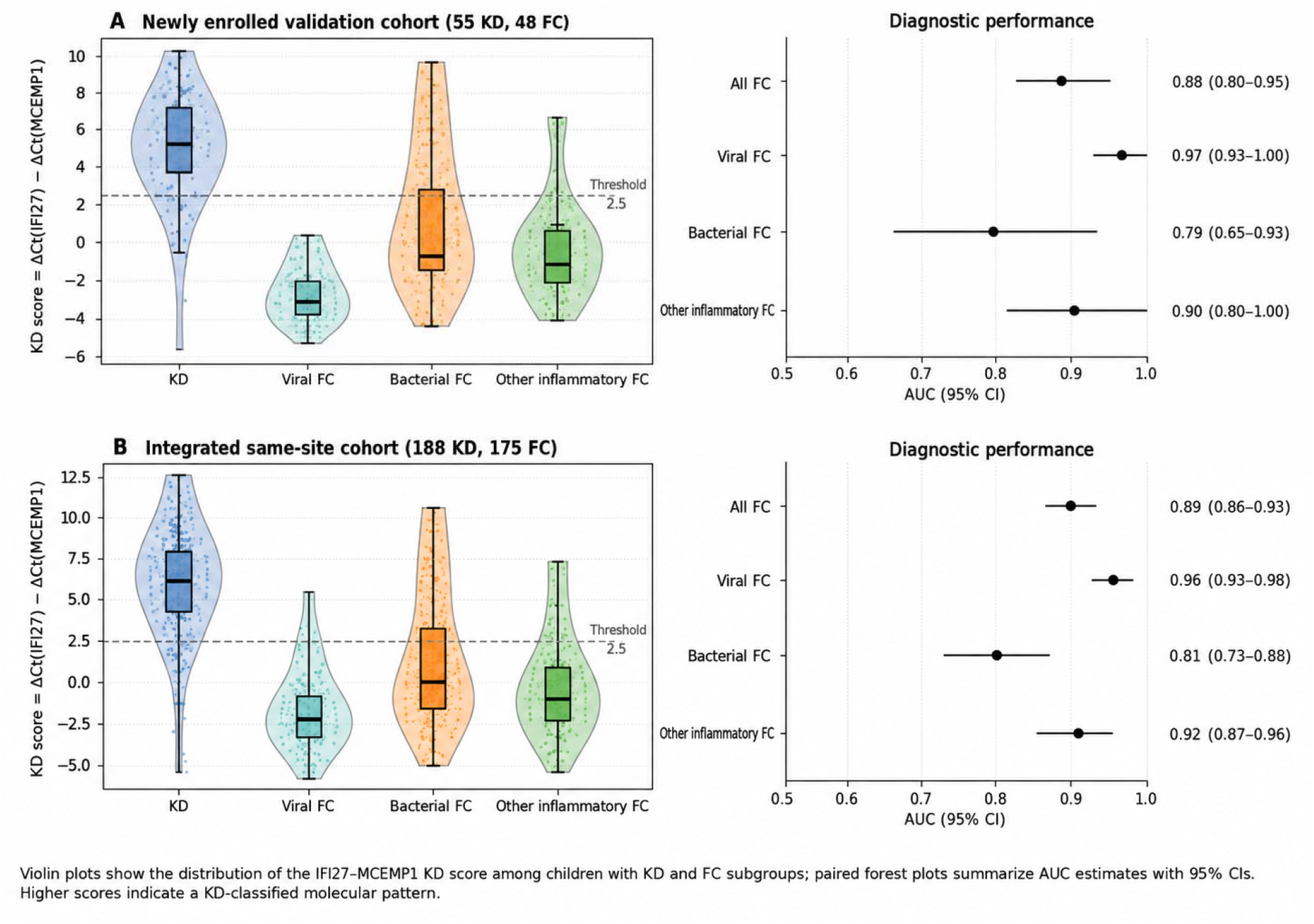
IFI27–MCEMP1 interferon–myeloid axis in Kawasaki disease and febrile controls. Violin plots show the distribution of the IFI27–MCEMP1 KD score among children with KD and FC subgroups, and paired receiver operating characteristic or forest-plot panels summarize AUC estimates with 95% CIs. **A**, Newly enrolled validation cohort comprising 55 KD and 48 FC participants. The IFI27–MCEMP1 score distinguished KD from all FC with an AUC of 0.88 (95% CI, 0.80–0.95). Comparator-specific AUCs were 0.97 (95% CI, 0.93–1.00) for viral FC, 0.79 (95% CI, 0.65–0.93) for bacterial FC, and 0.90 (95% CI, 0.79–1.00) for other inflammatory FC. **B**, Integrated same-site cohort comprising 188 KD and 175 FC participants. The IFI27–MCEMP1 score distinguished KD from all FC with an AUC of 0.89 (95% CI, 0.86–0.93). Comparator-specific AUCs were 0.96 (95% CI, 0.93–0.98) for viral FC, 0.81 (95% CI, 0.73–0.88) for bacterial FC, and 0.92 (95% CI, 0.87–0.96) for other inflammatory FC. Higher KD scores indicate a KD-classified molecular pattern. Error bars denote 95% CIs. **Abbreviations:** AUC, area under the receiver operating characteristic curve; FC, febrile control; KD, Kawasaki disease.

### Candidate immune-regulatory lncRNA selection

Three candidate lncRNAs were selected for evaluation: CHROMR, MALAT1, and NEAT1. These transcripts were chosen because of prior evidence linking them to immune transcriptional regulation, interferon-responsive programs, myeloid inflammatory signaling, or nuclear organization during inflammatory stress. CHROMR was evaluated as a candidate lncRNA related to interferon-stimulated gene regulation and macrophage-response biology. MALAT1 was evaluated because of its reported involvement in inflammatory signaling and transcriptional regulation in immune and vascular contexts. NEAT1 was evaluated because of its role in paraspeckle formation and interferon-responsive transcriptional organization.

The lncRNA analyses were designed to determine whether these candidate transcripts differed between KD and febrile controls and whether their expression patterns were associated with IFI27, MCEMP1, or the composite IFI27–MCEMP1 diagnostic score. These analyses were considered exploratory mechanistic-association analyses and were not designed to establish direct transcriptional regulation or causality.

### Immunologic and transcript-association outcomes

The primary diagnostic outcome was differential expression of CHROMR, MALAT1, and NEAT1 between KD and febrile controls; pairwise associations between lncRNA ΔCt values and IFI27 or MCEMP1 ΔCt values; and associations between candidate lncRNAs, clinical variables, laboratory variables, and the IFI27–MCEMP1 KD score. Differential-expression analyses are presented in Figure 3. Pairwise lncRNA–mRNA associations are presented in Figure 4. Univariate and multivariable analyses relating lncRNAs and clinical variables to the KD score are presented in Figure 5. A summary of lncRNA differential-expression and transcript-association findings is provided in **Table 5**. The integrated biological interpretation of these findings is shown schematically in **Figure 6**.

**Table 5.** Summary of lncRNA differential-expression and transcript-association findings in the integrated same-site cohort.

| Transcript | KD vs FC expression pattern | Association with IFI27 ΔCt | Association with MCEMP1 ΔCt | Biological interpretation |
| --- | --- | --- | --- | --- |
| CHROMR | Higher ΔCt in KD, indicating lower relative expression; Holm-adjusted P < .001 | Positive association; r = 0.55; Holm-adjusted P < .001 | Not significant after multiple-comparison adjustment | Strongest lncRNA correlate of the interferon-associated IFI27 component; consistent with coordinated attenuation of interferon-associated transcription |
| MALAT1 | Higher ΔCt in KD, indicating lower relative expression; Holm-adjusted P < .001 | Positive association; r = 0.15; Holm-adjusted P = .022 | Inverse association; r = -0.15; Holm-adjusted P = .022 | Weaker but directionally informative association with both interferon and myeloid components; may reflect broader perturbation of immune transcriptional regulation |
| NEAT1 | No significant differential-expression signal; Holm-adjusted P = .081 | Not significant after multiple-comparison adjustment | Not significant after multiple-comparison adjustment | No KD-specific differential-expression or transcript-association signal detected in this cohort |

**Abbreviations:** FC, febrile control; IFI27, interferon alpha-inducible protein 27; KD, Kawasaki disease; lncRNA, long noncoding RNA; MCEMP1, mast cell-expressed membrane protein 1.
**Note:** Analyses were performed in the integrated same-site cohort of 188 KD and 175 FC participants. Higher ΔCt values indicate lower relative expression. Differential-expression analyses used two-sided Mann–Whitney U tests with Holm adjustment across the three candidate lncRNAs. Pairwise transcript associations used Pearson correlations with Holm adjustment across six lncRNA–mRNA tests.

**Figure 3.**
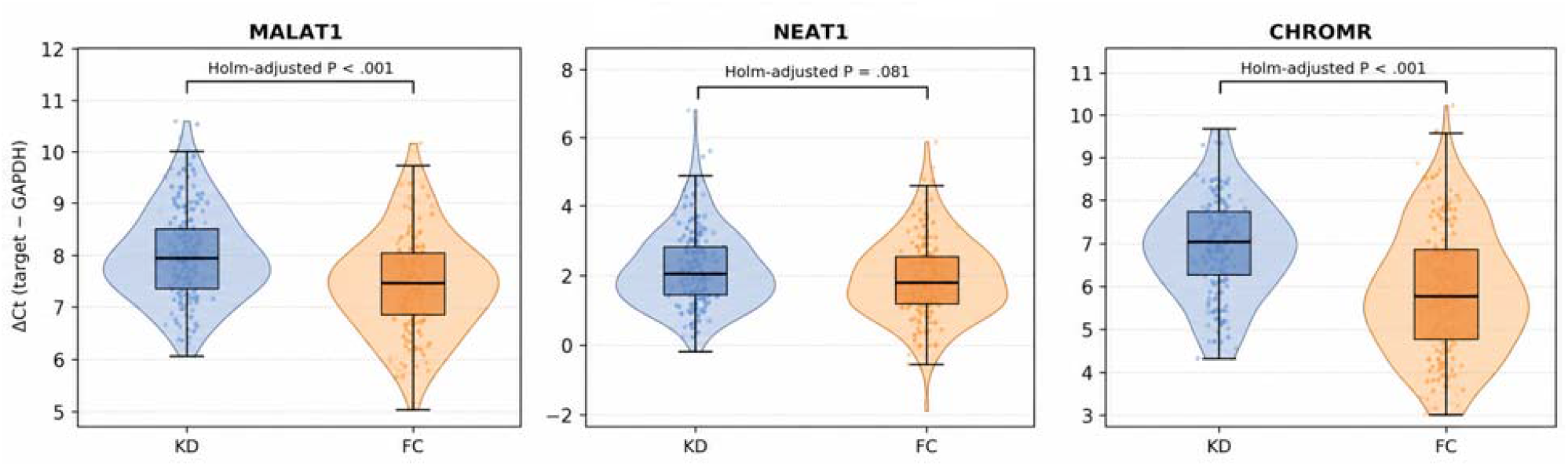
PBMC-derived CHROMR and MALAT1 are reduced in Kawasaki disease. Violin plots with embedded boxplots show ΔCt distributions for PBMC-derived CHROMR, MALAT1, and NEAT1 in children with KD and FC in the integrated same-site cohort of 188 KD and 175 FC participants. Higher ΔCt values indicate lower relative expression. Group comparisons used two-sided Mann–Whitney U tests with Holm adjustment across the three candidate lncRNAs. CHROMR and MALAT1 ΔCt values were significantly higher in KD than in FC, indicating lower relative expression in KD (Holm-adjusted P < .001 for both). NEAT1 did not differ significantly between KD and FC (Holm-adjusted P = .081). **Abbreviations:** FC, febrile control; KD, Kawasaki disease; lncRNA, long noncoding RNA; PBMC, peripheral blood mononuclear cell.

**Figure 4.**
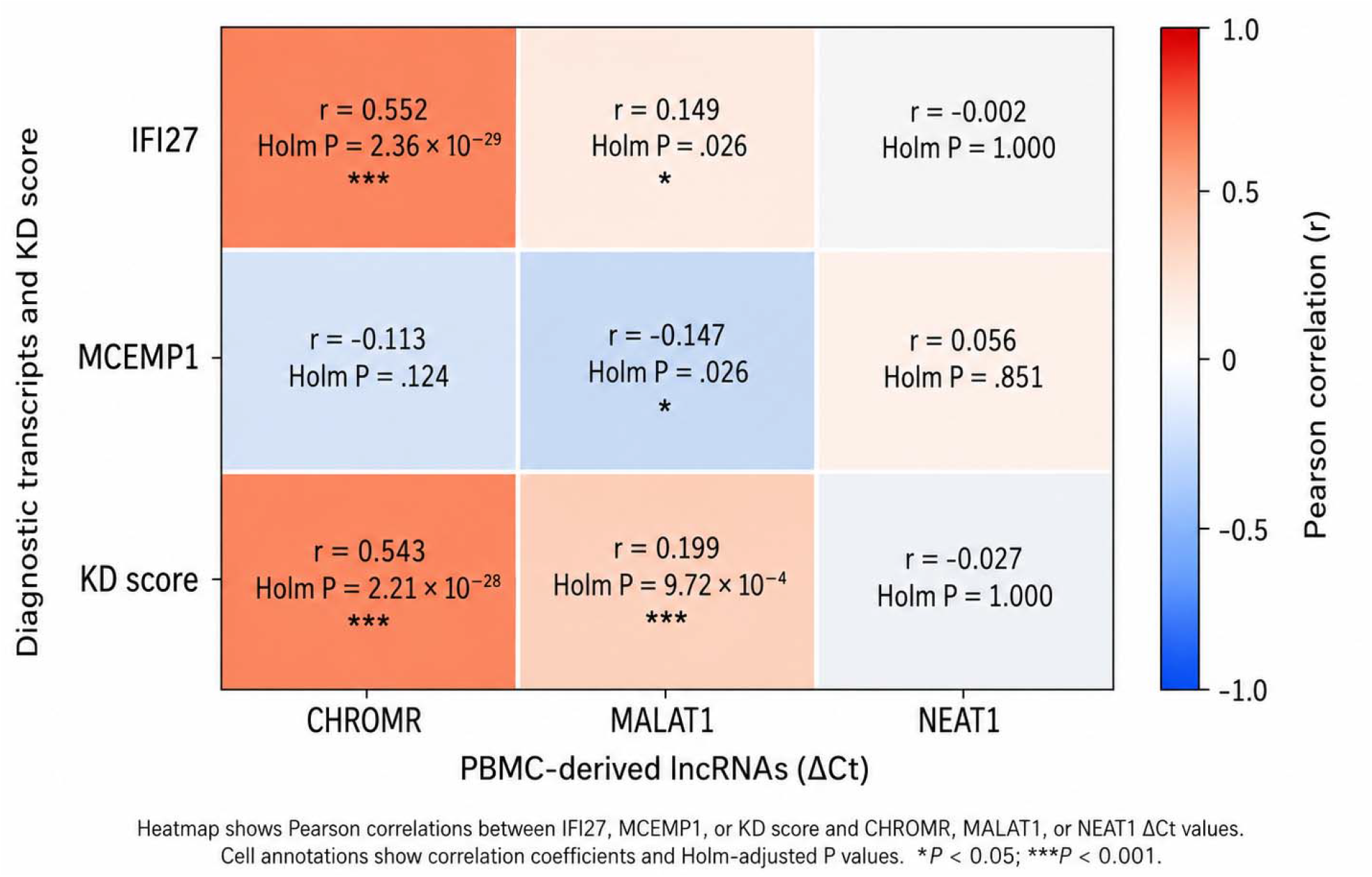
lncRNA associations with interferon and myeloid transcript components. Heatmap showing Pearson correlations between IFI27 or MCEMP1 ΔCt values and CHROMR, MALAT1, or NEAT1 ΔCt values across the integrated same-site cohort (n = 363). Cell annotations show correlation coefficients and Holm-adjusted P values across six pairwise lncRNA–mRNA tests. CHROMR showed the strongest positive association with IFI27 ΔCt (r = 0.55; adjusted P < .001). MALAT1 showed weaker associations with the two diagnostic transcripts, including a positive association with IFI27 ΔCt (r = 0.15; adjusted P = .022) and an inverse association with MCEMP1 ΔCt (r = −0.15; adjusted P = .022). NEAT1 was not significantly associated with either diagnostic transcript after multiple-comparison adjustment. **Abbreviations:** FC, febrile control; IFI27, interferon alpha-inducible protein 27; KD, Kawasaki disease; lncRNA, long noncoding RNA; MCEMP1, mast cell-expressed membrane protein 1; PBMC, peripheral blood mononuclear cell.

**Figure 5.**
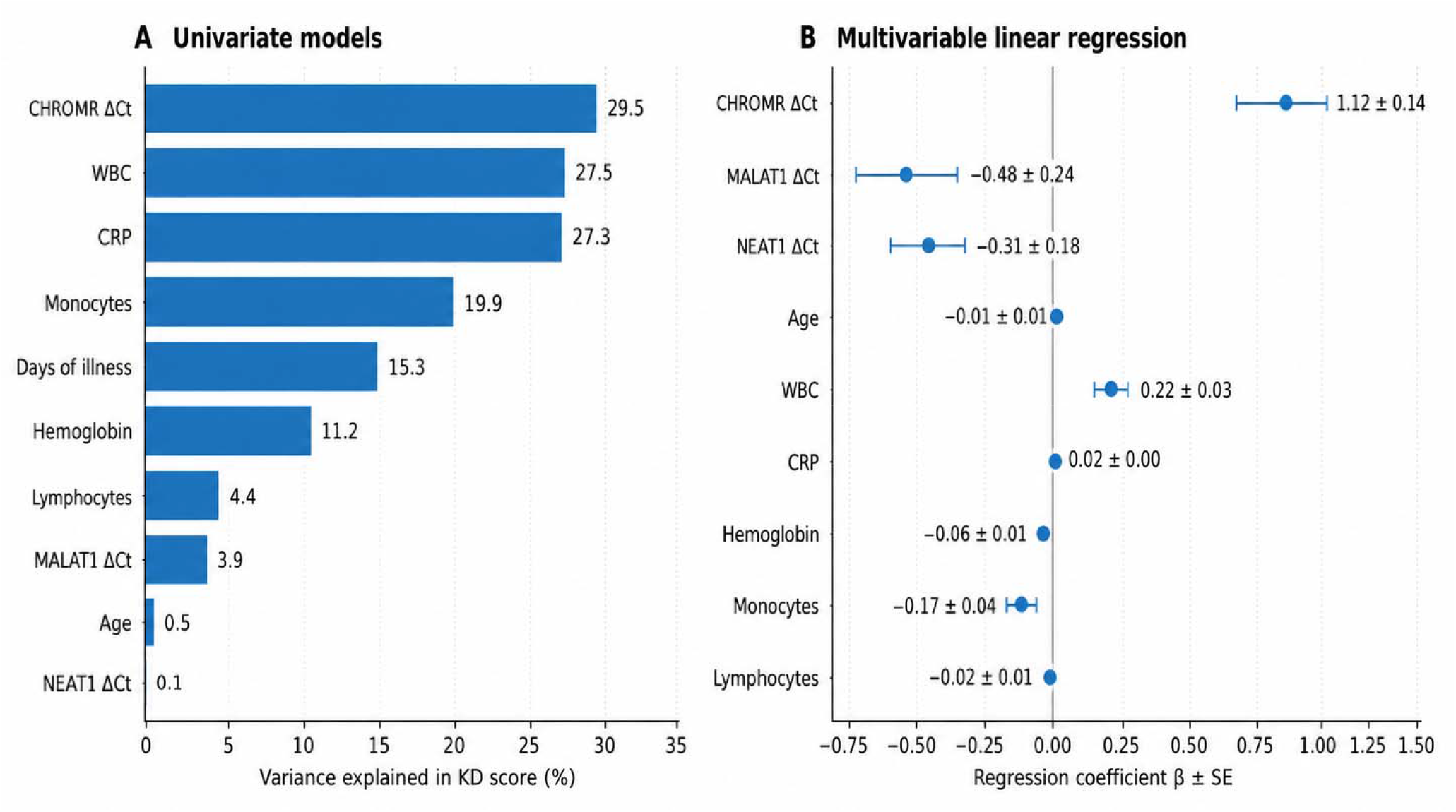
Clinical and lncRNA correlates of the IFI27–MCEMP1 immune axis. **A**, Percentage of IFI27–MCEMP1 KD-score variance explained by individual lncRNA and clinical variables in univariate linear models. **B**, Multivariable linear-regression coefficients with standard errors for lncRNAs and clinical covariates in relation to the IFI27–MCEMP1 KD score. Positive coefficients indicate a higher KD score with increasing predictor values; negative coefficients indicate a lower KD score with increasing predictor values. These analyses evaluate association with the diagnostic score and are not intended to establish incremental diagnostic utility, direct transcriptional regulation, or causal relationships. **Abbreviations:** FC, febrile control; IFI27, interferon alpha-inducible protein 27; KD, Kawasaki disease; lncRNA, long noncoding RNA; MCEMP1, mast cell-expressed membrane protein 1.

**Figure 6.**
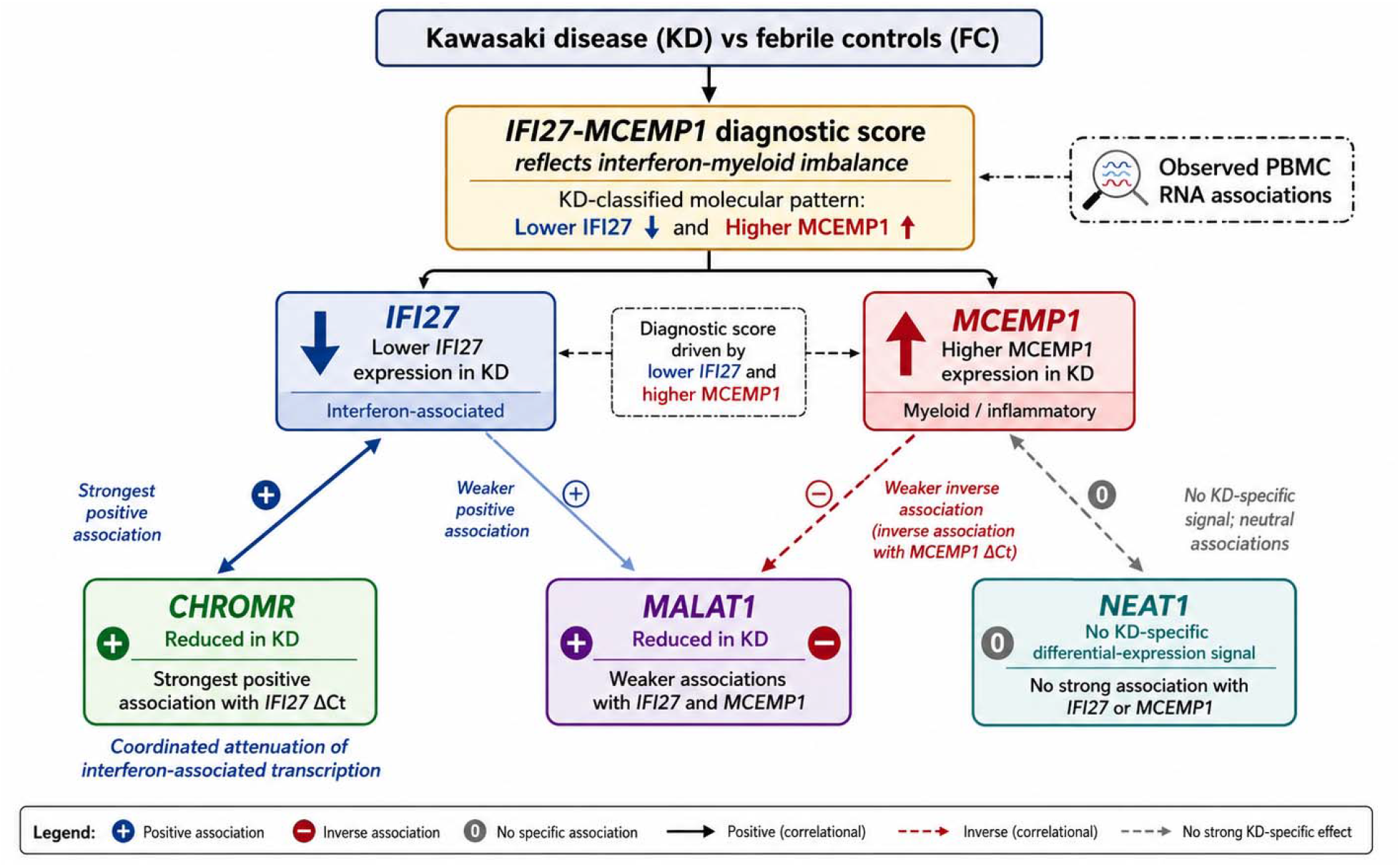
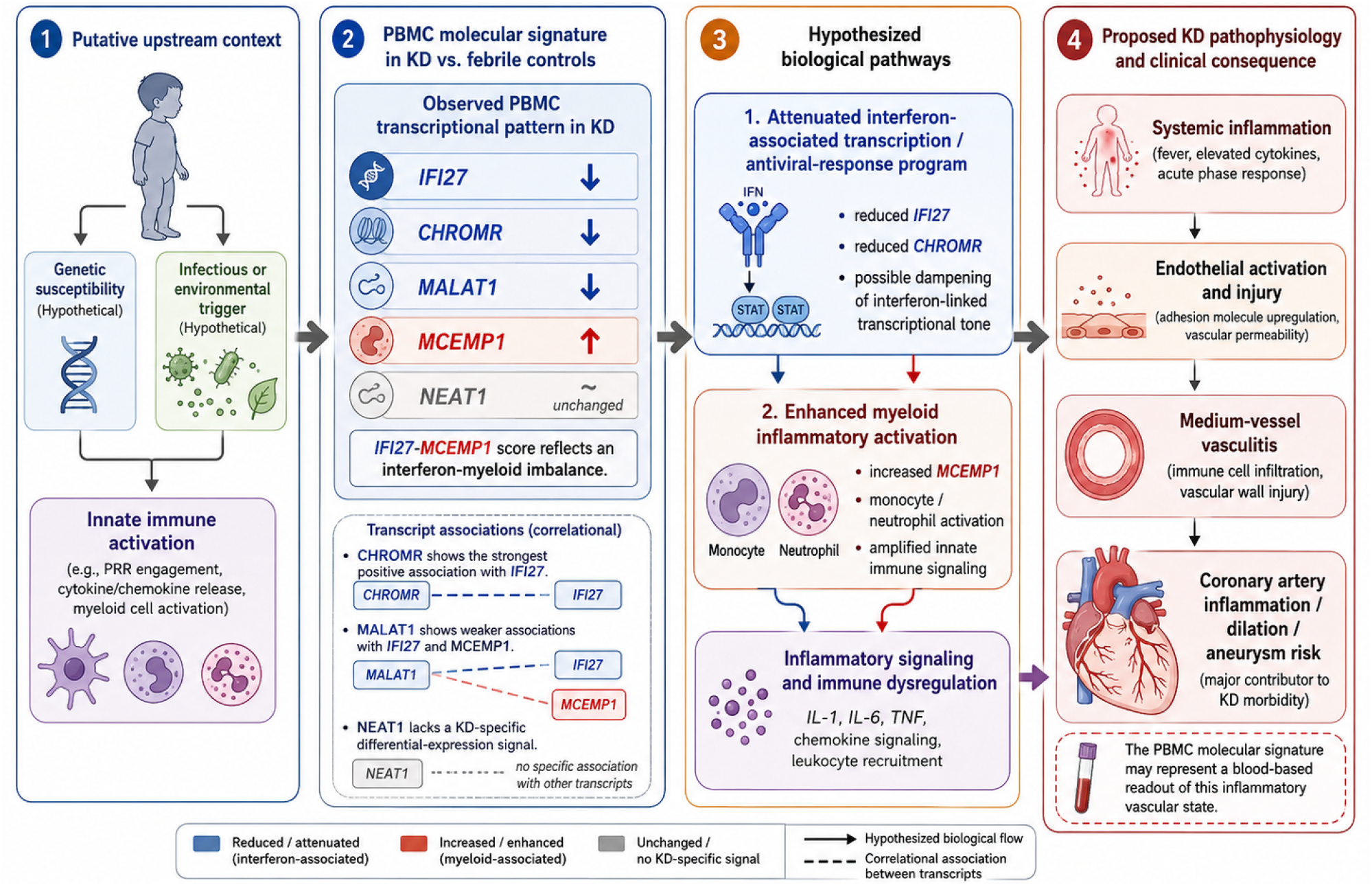
Proposed lncRNA-associated interferon–myeloid model of Kawasaki disease vasculitis. **A**, Schematic model summarizing observed PBMC RNA associations in KD. The IFI27– MCEMP1 diagnostic score reflects an interferon–myeloid imbalance, with lower IFI27 expression and higher MCEMP1 expression in KD relative to febrile controls. CHROMR was reduced in KD and showed the strongest positive association with IFI27 ΔCt, consistent with coordinated attenuation of interferon-associated transcription. MALAT1 was reduced in KD and showed weaker associations with IFI27 and MCEMP1, including an inverse association with MCEMP1 ΔCt. NEAT1 did not show a KD-specific differential-expression signal. All lncRNA–mRNA relationships are correlational and do not establish direct regulation. **B**, Hypothesized biological pathway model linking the observed PBMC RNA signature to KD immune dysregulation and vascular injury. In KD relative to febrile controls, lower IFI27 expression together with lower CHROMR and MALAT1 expression, and higher MCEMP1 expression, are hypothesized to reflect an interferon–myeloid imbalance. Reduced IFI27 and CHROMR are consistent with attenuation of interferon-associated or antiviral-response transcriptional programs, whereas increased MCEMP1 is consistent with enhanced myeloid inflammatory activation. MALAT1 may reflect broader perturbation of immune transcriptional regulation, while NEAT1 did not demonstrate a KD-specific signal. These molecular changes may converge on inflammatory signaling, endothelial activation, medium-vessel vasculitis, and coronary artery inflammation, dilation, or aneurysm risk. The PBMC molecular signature may therefore represent a blood-based readout of this inflammatory vascular state. This model is hypothetical and intended for biological interpretation; transcript associations are correlational and do not establish direct regulation or causality. **Abbreviations:** FC, febrile control; IFI27, interferon alpha-inducible protein 27; KD, Kawasaki disease; lncRNA, long noncoding RNA; MCEMP1, mast cell-expressed membrane protein 1; PBMC, peripheral blood mononuclear cell; RT-qPCR, reverse transcription quantitative polymerase chain reaction.

Secondary outcomes included performance of the IFI27–MCEMP1 KD score for distinguishing KD from febrile controls in the newly enrolled validation cohort. Diagnostic performance was assessed overall and by febrile-control subgroup. Diagnostic performance was also evaluated in the integrated same-site cohort to confirm reproducibility across all available same-site samples.

### Statistical analysis

Continuous variables were summarized as means with standard deviations or medians with interquartile ranges, depending on distribution. Categorical variables were summarized as counts and percentages. Between-group comparisons were performed using t tests for approximately normally distributed continuous variables, Wilcoxon rank-sum or Mann–Whitney U tests for skewed continuous variables, and χ^2^ or Fisher exact tests for categorical variables, as appropriate.

Diagnostic performance of the IFI27–MCEMP1 KD score was evaluated using receiver operating characteristic curve analysis. Area under the receiver operating characteristic curve values and 95% confidence intervals were estimated using the DeLong method. Sensitivity, specificity, positive predictive value, and negative predictive value were calculated at the prespecified diagnostic-positive threshold of KD score ≥2.5. Exact binomial methods were used to estimate confidence intervals for sensitivity, specificity, positive predictive value, and negative predictive value. Comparator-specific diagnostic analyses were performed by comparing KD cases with viral, bacterial, and other inflammatory febrile-control subgroups. Predictive values were reported for the full febrile-control comparison because they depend on the case-control composition and may not generalize to clinical prevalence.

Differential lncRNA expression between KD and febrile controls was assessed using ΔCt values, with higher ΔCt values interpreted as lower relative expression. Group comparisons for CHROMR, MALAT1, and NEAT1 were performed using two-sided Mann–Whitney U tests, with Holm adjustment for multiple comparisons across the three candidate lncRNAs.

Pairwise transcript associations were evaluated using Pearson correlation coefficients between lncRNA ΔCt values and IFI27 or MCEMP1 ΔCt values. Six lncRNA–mRNA correlations were tested, and P values were adjusted using the Holm method. Correlation analyses were performed in the integrated same-site cohort.

Linear regression models were used to evaluate relationships between candidate lncRNAs, clinical variables, laboratory variables, and the IFI27–MCEMP1 KD score. In univariate models, each lncRNA or clinical variable was evaluated separately to estimate the percentage of KD-score variance explained. Multivariable linear regression was then used to assess associations between lncRNAs and the KD score after adjustment for selected clinical and laboratory covariates, including age, white blood cell count, C-reactive protein, hemoglobin, and leukocyte differential counts when available. Regression coefficients are reported with standard errors. These models were used to evaluate association with the diagnostic score and were not interpreted as evidence of incremental diagnostic utility, direct transcriptional regulation, causal mediation, or disease mechanism.

Analyses were performed using complete cases without imputation. All statistical tests were two-sided, and P values less than .05 were considered statistically significant after adjustment where applicable. Statistical analyses were performed using R version 4.3.

### Reporting standards and assay reproducibility

The diagnostic-validation component of the study was reported in accordance with the Standards for Reporting Diagnostic Accuracy Studies guideline. The RT-qPCR assay-reporting components followed the Minimum Information for Publication of Quantitative Real-Time PCR Experiments recommendations.

## RESULTS

### Study population and PBMC RNA profiling cohorts

The study included a newly enrolled cohort and two previously characterized same-site cohorts used for integrated peripheral blood mononuclear cell (PBMC) RNA analyses. The newly enrolled cohort, designated batch 3, included 103 children evaluated for suspected Kawasaki disease (KD) or alternative febrile illnesses, comprising 55 children with KD and 48 febrile controls. Two previously characterized same-site cohorts were integrated for transcript-association analyses: batch 1 included 53 children with KD and 81 febrile controls, and batch 2 included 80 children with KD and 46 febrile controls. Integration of all three batches yielded an analytic cohort of 188 children with KD and 175 febrile controls, for a total of 363 participants available for PBMC long noncoding RNA (lncRNA), mRNA, and IFI27–MCEMP1 axis analyses (Table 1; Figure 1).

In the newly enrolled cohort, children with KD were younger than febrile controls, with a median age of 2.25 years compared with 3.79 years in febrile controls (P < 0.001). Sex distribution was similar between groups, with male children comprising 69.1% of the KD group and 66.7% of the febrile-control group (P = 0.793). Fever duration at sample collection was longer in KD than in febrile controls, with median durations of 6 days and 2 days, respectively (P < 0.001). Children with KD also had lower hemoglobin concentrations, higher C-reactive protein concentrations, and higher white blood cell counts than febrile controls (all P < 0.001). Platelet counts did not differ significantly between groups (P = 0.070). Among the 55 children with KD, 21 (38.2%) had incomplete KD, and 19 (34.5%) had coronary artery dilation or aneurysm. Among febrile controls, 15 (31.3%) had viral febrile illness, 20 (41.7%) had bacterial febrile illness, and 13 (27.1%) had other inflammatory febrile illness (Table 2).

### Reproducibility of the IFI27–MCEMP1 interferon–myeloid axis

We first evaluated whether the IFI27–MCEMP1 transcript axis, previously validated as a two-gene KD-associated molecular score, reproduced the expected separation between KD and febrile controls in the newly enrolled cohort. The score is defined as ΔCt(IFI27) − ΔCt(MCEMP1), where higher ΔCt indicates lower relative expression. Thus, higher scores reflect the KD-associated pattern of relatively lower IFI27 expression and higher MCEMP1 expression.

In the newly enrolled cohort, the IFI27–MCEMP1 axis distinguished KD from all febrile controls with an area under the receiver operating characteristic curve (AUC) of 0.88 (95% CI, 0.80–0.95). At the prespecified operating point of KD score ≥2.5, sensitivity was 81.8% (95% CI, 69.1–90.9), specificity was 87.5% (95% CI, 74.8–95.3), positive predictive value was 88.2% (95% CI, 76.1–95.6), and negative predictive value was 80.8% (95% CI, 67.5–90.4) (Table 4; Figure 2A).

The IFI27–MCEMP1 axis showed comparator-dependent discrimination across febrile-control subgroups. In the newly enrolled cohort, separation was strongest against viral febrile controls, with an AUC of 0.97 (95% CI, 0.93–1.00) and specificity of 100.0% (95% CI, 78.2–100.0). Discrimination was lower against bacterial febrile controls, with an AUC of 0.79 (95% CI, 0.65–0.93) and specificity of 75.0% (95% CI, 50.9–91.3). Against other inflammatory febrile controls, the AUC was 0.90 (95% CI, 0.79–1.00), with specificity of 92.3% (95% CI, 64.0–99.8) (Table 4; Figure 2A).

In the integrated same-site cohort, the IFI27–MCEMP1 axis showed similar overall performance. The score distinguished KD from all febrile controls with an AUC of 0.89 (95% CI, 0.86–0.93). At the same prespecified threshold, sensitivity was 85.6% (95% CI, 79.8–90.3), specificity was 84.6% (95% CI, 78.4–89.6), positive predictive value was 85.6% (95% CI, 79.8–90.3), and negative predictive value was 84.6% (95% CI, 78.4– 89.6) (Table 4; Figure 2B). Comparator-specific discrimination in the integrated cohort was again highest against viral febrile controls (AUC, 0.96; 95% CI, 0.93–0.98), followed by other inflammatory febrile controls (AUC, 0.92; 95% CI, 0.87–0.96) and bacterial febrile controls (AUC, 0.81; 95% CI, 0.73–0.88) (Table 4; Figure 2B). These findings supported the reproducibility of the IFI27–MCEMP1 axis as a PBMC immune-transcript pattern characterized by relative interferon attenuation and myeloid activation in KD.

### PBMC-derived CHROMR and MALAT1 are reduced in Kawasaki disease

We next evaluated whether candidate immune-regulatory lncRNAs differed between KD and febrile controls in the integrated same-site cohort. Expression of CHROMR, MALAT1, and NEAT1 was measured by RT-qPCR and normalized to GAPDH using ΔCt values. Primer sequences, amplicon sizes, amplification efficiencies, and linearity metrics for the lncRNA RT-qPCR assays are shown in Table 3.

CHROMR ΔCt values were significantly higher in KD than in febrile controls, indicating lower relative CHROMR expression in KD (Holm-adjusted P < 0.001). MALAT1 showed a similar pattern, with significantly higher ΔCt values in KD than in febrile controls, also indicating lower relative expression in KD (Holm-adjusted P < 0.001). In contrast, NEAT1 did not show a statistically significant difference between KD and febrile controls after multiple-comparison adjustment (Holm-adjusted P = 0.081) (Figure 3; Table 5). These findings indicate that CHROMR and MALAT1, but not NEAT1, are altered in the PBMC transcriptome of children with KD relative to febrile controls.

### CHROMR is strongly associated with the IFI27 interferon component

We then assessed whether candidate lncRNAs were associated with the mRNA components of the IFI27–MCEMP1 axis. Across the integrated same-site cohort, CHROMR showed the strongest transcript association. CHROMR ΔCt was positively correlated with IFI27 ΔCt (r = 0.55; Holm-adjusted P < 0.001) (Figure 4; Table 5). Because higher ΔCt values indicate lower relative expression, this positive association indicates that lower CHROMR expression tracked with lower IFI27 expression across the cohort.

This relationship was specific to the interferon-associated component of the axis. CHROMR did not show a significant association with MCEMP1 after multiple-comparison adjustment. Thus, among the candidate lncRNAs evaluated, CHROMR was the dominant correlate of the IFI27 component, consistent with coordinated attenuation of CHROMR and interferon-associated transcription in KD.

### MALAT1 shows weaker associations with both interferon and myeloid components

MALAT1 showed weaker but statistically significant associations with both components of the IFI27–MCEMP1 axis. MALAT1 ΔCt was positively associated with IFI27 ΔCt (r = 0.15; Holm-adjusted P = 0.022), indicating that lower MALAT1 expression was modestly associated with lower IFI27 expression. MALAT1 ΔCt was inversely associated with MCEMP1 ΔCt (r = −0.15; Holm-adjusted P = 0.022), indicating that lower MALAT1 expression was modestly associated with higher MCEMP1 expression (Figure 4; Table 5).

This pattern aligned directionally with the KD-associated IFI27–MCEMP1 axis: reduced MALAT1 expression was associated with both reduced IFI27 and increased MCEMP1. Although the magnitude of these associations was modest, the bidirectional relationship with both mRNA components suggests that MALAT1 may reflect a broader PBMC immune-transcriptional state linking interferon attenuation and myeloid activation.

### NEAT1 does not show a KD-specific transcript association signal

Unlike CHROMR and MALAT1, NEAT1 did not show a significant KD-specific differential-expression signal after multiple-comparison adjustment and was not significantly associated with either IFI27 or MCEMP1 after adjustment across the six lncRNA–mRNA tests (Figure 4; Table 5). This negative finding suggests that the observed lncRNA associations were not explained by a generalized shift in all inflammation-related lncRNAs. Instead, the pattern was selective for CHROMR and MALAT1 within the candidate lncRNA panel evaluated.

### Clinical and lncRNA correlates of the IFI27–MCEMP1 axis

Univariate linear models were used to estimate the proportion of IFI27–MCEMP1 axis variance explained by individual lncRNA and clinical variables. These analyses evaluated whether candidate lncRNAs and clinical parameters were associated with the composite IFI27–MCEMP1 score, rather than whether they improved diagnostic classification beyond the two-transcript axis. The variance-explained analysis is summarized in Figure 5A.

Multivariable linear regression was then used to evaluate associations between lncRNAs and the IFI27–MCEMP1 axis after adjustment for clinical and laboratory covariates. Regression coefficients and standard errors are shown in Figure 5B. These analyses supported an association between selected lncRNAs and the IFI27–MCEMP1 immune axis while preserving the distinction between statistical association and direct regulatory or causal inference. The models were not interpreted as evidence of direct transcriptional regulation, causal mediation, or incremental diagnostic utility (Figure 5).

### Integrated lncRNA-associated interferon–myeloid model of Kawasaki disease

Together, the transcript patterns supported an integrated PBMC RNA model in which KD is characterized by an lncRNA-associated interferon–myeloid imbalance. In this model, KD is marked by lower IFI27 expression and higher MCEMP1 expression relative to febrile controls. Among the candidate lncRNAs, CHROMR and MALAT1 were lower in KD, whereas NEAT1 did not show a KD-specific signal. CHROMR showed the strongest association with the IFI27 interferon-associated component, while MALAT1 showed weaker associations with both IFI27 and MCEMP1, including an inverse association with MCEMP1 ΔCt (Table 5; Figure 6A).

These observations were summarized in a hypothesized biological pathway model linking PBMC RNA perturbation to KD immune dysregulation and vascular inflammation. The model proposes that reduced IFI27, CHROMR, and MALAT1 expression, together with increased MCEMP1 expression, may reflect attenuation of interferon-associated transcriptional programs coupled with enhanced myeloid inflammatory activation. This PBMC RNA architecture may provide a circulating immune readout of inflammatory signaling, endothelial activation, medium-vessel vasculitis, and coronary artery involvement in KD. The model remains hypothesis-generating, and the observed lncRNA–mRNA relationships do not establish direct regulation or causality (Figure 6B).

## DISCUSSION

In this PBMC transcript association study, we evaluated whether candidate long noncoding RNAs (lncRNAs) are linked to an IFI27–MCEMP1 interferon–myeloid transcriptomic axis in Kawasaki disease (KD). Three principal findings emerged. First, the IFI27–MCEMP1 axis reproduced the expected KD-associated immune-transcript pattern in a newly enrolled cohort and in the integrated same-site cohort, with relative attenuation of the interferon-associated transcript IFI27 and elevation of the myeloid-associated transcript MCEMP1 in KD compared with febrile controls. Second, two candidate immune-regulatory lncRNAs, CHROMR and MALAT1, showed lower relative expression in KD, whereas NEAT1 did not show a significant KD-specific differential-expression signal. Third, CHROMR demonstrated the strongest association with the IFI27 interferon component, while MALAT1 showed weaker but directionally informative associations with both IFI27 and MCEMP1. Together, these findings support an lncRNA-associated interferon–myeloid RNA architecture in KD and generate testable hypotheses regarding immune transcriptional regulation in KD vasculitis.

KD remains an enigmatic pediatric vasculitis in which clinical inflammation, immune activation, and vascular injury are closely linked but incompletely understood. The acute illness involves systemic inflammatory responses, endothelial activation, and coronary artery susceptibility, yet the circulating immune programs that distinguish KD from other febrile illnesses remain difficult to define. In this context, the IFI27–MCEMP1 axis provides a useful immunologic anchor. IFI27 is an interferon-stimulated gene associated with antiviral-response biology, whereas MCEMP1 is linked to myeloid inflammatory activation. The combination of lower IFI27 and higher MCEMP1 expression in KD suggests that the disease is not simply characterized by global immune activation, but instead by a distinct pattern of interferon attenuation coupled with myeloid activation. The present study extends this concept by showing that selected lncRNAs, particularly CHROMR and MALAT1, are associated with this immune-transcript configuration.

The reproducibility of the IFI27–MCEMP1 axis across the newly enrolled and integrated cohorts is important for interpreting the lncRNA findings. The score distinguished KD from febrile controls with similar discrimination in both datasets, and the comparator-specific pattern was biologically plausible: separation was strongest against viral febrile controls and lower against bacterial febrile controls. This pattern is consistent with the possibility that viral illnesses maintain a stronger interferon-associated transcriptional state, whereas bacterial infections and KD may share greater overlap in myeloid or neutrophil-associated inflammation. Therefore, the IFI27–MCEMP1 axis may be best understood not merely as a diagnostic classifier, but as a compressed representation of a broader host-response contrast between interferon-dominant and myeloid-dominant inflammatory states.

The strongest lncRNA association observed in this study was between CHROMR and IFI27. Because higher ΔCt values indicate lower relative expression, the positive correlation between CHROMR ΔCt and IFI27 ΔCt indicates that reduced CHROMR expression tracked with reduced IFI27 expression across the integrated cohort. This finding is consistent with coordinated attenuation of CHROMR and interferon-associated transcription in KD. CHROMR has been implicated in macrophage immune responses and interferon-stimulated gene regulation, including interactions with STAT1- and IRF1-associated transcriptional programs [10]. The observed CHROMR–IFI27 relationship therefore provides a biologically plausible link between a candidate lncRNA and the interferon-associated component of the KD transcriptomic axis. However, the current data cannot determine whether CHROMR directly regulates IFI27, whether both transcripts are downstream of common upstream signaling, or whether their correlation reflects differences in PBMC cellular composition.

MALAT1 showed a broader but weaker pattern of association. MALAT1 expression was lower in KD and was modestly associated with both sides of the IFI27–MCEMP1 axis: positively with IFI27 ΔCt and inversely with MCEMP1 ΔCt. Interpreted in expression terms, lower MALAT1 expression was associated with lower IFI27 and higher MCEMP1, matching the directionality of the KD-associated interferon–myeloid pattern. MALAT1 has been linked to inflammatory transcriptional regulation, including pathways involving NF-κB, immune-cell activation, and endothelial biology [12]. Its association with both IFI27 and MCEMP1 suggests that MALAT1 may reflect a broader PBMC transcriptional state rather than a specific interferon-only program. This raises the possibility that MALAT1 perturbation may mark an interface between interferon attenuation, myeloid activation, and vascular inflammatory responses in KD. Because the observed correlations were modest, this interpretation should be considered hypothesis-generating rather than mechanistically definitive.

In contrast, NEAT1 did not show a statistically significant differential-expression signal in KD and was not significantly associated with either IFI27 or MCEMP1 after multiple-comparison adjustment. This negative finding is useful. NEAT1 is involved in paraspeckle formation and has been implicated in inflammatory and interferon-responsive transcriptional organization [11], but its lack of a clear signal in this cohort suggests that the lncRNA alterations observed in KD are not simply a nonspecific consequence of acute inflammation or global lncRNA dysregulation. Instead, the differential pattern across CHROMR, MALAT1, and NEAT1 supports a degree of transcript specificity within the candidate lncRNA panel.

These observations support a conceptual model in which KD is characterized by a PBMC RNA state involving interferon attenuation, myeloid activation, and selective lncRNA perturbation. In this model, lower IFI27 and CHROMR expression may reflect remodeling or suppression of interferon-associated antiviral-response programs, whereas higher MCEMP1 expression reflects enhanced myeloid inflammatory activation. MALAT1 may mark a broader immune-transcriptional perturbation that bridges these two components. Such a model is compatible with current views of KD as an immune-mediated vasculitis in which innate immune activation, endothelial inflammation, and coronary artery injury emerge from dysregulated host responses. Importantly, the PBMC transcriptomic pattern measured here should be interpreted as a circulating immune readout. It does not directly measure immune activity within the coronary arterial wall, nor does it establish the tissue-level mechanisms of coronary arteritis.

The immunologic implications of an interferon–myeloid imbalance in KD are notable. Interferon-stimulated genes often dominate host responses to viral infection, whereas myeloid activation, neutrophil recruitment, macrophage activation, and inflammatory cytokine production are prominent features of vasculitic and autoinflammatory states. A KD-associated pattern of reduced IFI27 with increased MCEMP1 may therefore reflect an immune trajectory distinct from conventional antiviral febrile illness. The addition of lncRNA data suggests that this trajectory may be accompanied by altered regulatory RNA programs. If confirmed, such lncRNA-associated immune states could help define biologically meaningful KD endotypes, identify children at risk for persistent inflammation or coronary involvement, or clarify why some febrile inflammatory illnesses mimic KD clinically but differ molecularly.

The clinical relevance of these findings lies in the connection between immune interpretability and molecular phenotyping. A two-transcript score can be clinically useful, but mechanistic confidence increases when that score is embedded within a coherent immune network. By linking the IFI27–MCEMP1 axis to CHROMR and MALAT1, this study suggests that the KD-associated transcript pattern reflects a broader RNA architecture rather than an isolated biomarker pair. This is particularly relevant for future studies that aim to move beyond binary disease classification toward molecular stratification of KD. For example, lncRNA-associated interferon–myeloid patterns could be evaluated in relation to fever duration, coronary artery status, IVIG responsiveness, inflammatory persistence, or longitudinal treatment response. This study has several strengths. The analysis used a newly enrolled cohort and an integrated same-site cohort with consistent PBMC-based RT-qPCR profiling. The IFI27–MCEMP1 axis was evaluated using a prespecified score and threshold, reducing the risk of overfitting. The study included febrile controls with viral, bacterial, and other inflammatory illnesses, which is important because these are the clinical contexts in which KD immune signatures must be distinguished from overlapping inflammatory states. The lncRNA analyses were linked to a reproducible immune-transcript axis rather than conducted as isolated candidate comparisons, enabling a more coherent interpretation of the RNA findings.

Several limitations should be considered. First, the study is observational and based on transcript associations. It cannot establish direct lncRNA regulation of IFI27 or MCEMP1, causal mediation, or mechanistic control of KD inflammation. Functional perturbation experiments in relevant immune-cell models will be needed to determine whether CHROMR or MALAT1 directly modulate interferon-stimulated gene programs, myeloid activation, or endothelial-inflammatory pathways. Second, RNA was measured in PBMCs, which provide an accessible circulating immune compartment but do not capture granulocyte-rich whole-blood biology, vascular-wall immune activity, or tissue-resident inflammatory processes. Differences in PBMC subset composition may contribute to the observed transcript patterns. Third, the integrated cohort was derived from a single institution, and validation in additional geographic, ethnic, and clinical settings is needed. Fourth, febrile-control annotations in the historical integrated cohort included non-mutually exclusive categories for some participants; therefore, comparator-specific analyses should be interpreted as immune-subgroup contrasts rather than prevalence estimates. Fifth, the candidate lncRNAs were selected based on biological plausibility rather than unbiased transcriptome-wide discovery. Broader RNA-sequencing studies may identify additional lncRNA, enhancer RNA, circular RNA, or small RNA programs relevant to KD.

Future work should extend these findings in several directions. Longitudinal sampling before and after IVIG treatment could determine whether CHROMR, MALAT1, IFI27, and MCEMP1 normalize with clinical improvement or persist in children with ongoing vascular risk. Cell-type-resolved studies, including single-cell RNA sequencing or sorted immune-cell profiling, could define whether the observed lncRNA patterns arise from monocytes, lymphocytes, or shifts in PBMC composition. Functional studies could test whether CHROMR and MALAT1 influence interferon signaling, MCEMP1-associated myeloid activation, cytokine production, or endothelial activation in KD-relevant immune-cell systems. Finally, integration of lncRNA profiles with coronary artery phenotypes, IVIG response, and inflammatory trajectories could clarify whether this interferon–myeloid architecture defines clinically meaningful KD endotypes. In conclusion, PBMC-derived CHROMR and MALAT1 were reduced in KD and associated with components of the IFI27–MCEMP1 interferon–myeloid transcriptomic axis, whereas NEAT1 showed no clear KD-specific signal. These findings support a model in which KD is characterized by selective lncRNA perturbation accompanying interferon attenuation and myeloid activation. The results provide an interpretable PBMC RNA framework for KD immune dysregulation and generate mechanistic hypotheses for future studies of lncRNA regulation, vascular inflammation, and molecular endotyping in KD vasculitis.

## Data Availability

Individual-level participant data are not publicly available because of patient privacy protections and institutional ethical approvals at the Children's Hospital of Fudan University. Deidentified summary data, analytic code, and materials necessary to reproduce the reported analyses will be made available upon publication at the project repository.
Additional deidentified data may be considered through collaboration with the Children's Hospital of Fudan University, subject to institutional review and approval.

## Abbreviations

AUC: area under the receiver operating characteristic curve
Ct: cycle threshold
FC: febrile control
IFI27: interferon alpha-inducible protein 27
IVIG: intravenous immunoglobulin
KD: Kawasaki disease
lncRNA: long noncoding RNA
MALAT1: metastasis-associated lung adenocarcinoma transcript 1
MCEMP1: mast cell–expressed membrane protein 1
MIQE: Minimum Information for Publication of Quantitative Real-Time PCR Experiments
NEAT1: nuclear enriched abundant transcript 1
NPV: negative predictive value
PBMC: peripheral blood mononuclear cell
PPV: positive predictive value
qPCR: quantitative polymerase chain reaction
ROC: receiver operating characteristic
RT-qPCR: reverse transcription quantitative polymerase chain reaction
STARD: Standards for Reporting Diagnostic Accuracy Studies

